# Mapping Topic Change in Influential Hepatocellular Carcinoma Research: A Two-Cohort Bibliometric Analysis

**DOI:** 10.64898/2026.07.07.26357427

**Authors:** Zheng Su, Tinsley Li

**Affiliations:** AnyHelix Ltd., The Cloud, Tai Kok Tsui, Hong Kong, China; Computational Biology Group, FAB Tech, Yantian, Shenzhen, Guangdong, 518083, China

## Abstract

The therapeutic landscape for hepatocellular carcinoma (HCC) is evolving rapidly, necessitating scalable approaches to synthesize the expanding scientific literature. We characterized thematic shifts in HCC treatment and prognosis research by conducting a retrospective bibliometric analysis of influential publications from 2023 and 2024. Using the OpenAlex database, we identified the 50 most highly cited papers from each year based on eighteen-month post-publication citation counts. Large language models were deployed to extract, normalize, and classify concepts from unstructured text into canonical topics and parent themes, enabling quantitative year-over-year frequency comparisons. Analysis of these 100 papers revealed a distinct maturation in research focus. Although broad categories like general immunotherapy remained prevalent, their relative frequency declined in favor of specific dual immune checkpoint regimens, notably CTLA-4 inhibition and the durvalumab plus tremelimumab combination. Concurrently, parent themes related to radiomics, imaging, and health systems exhibited significant growth in the 2024 cohort. These findings demonstrate a thematic transition in high-impact HCC research from foundational immuno-oncology toward optimized combination therapies and precision diagnostics. Furthermore, this study highlights the utility of artificial intelligence-driven bibliometrics for objectively tracking dynamic conceptual shifts in oncology. A web interface for exploring the data is available at https://pri.pepkio.com/.

## Introduction

Hepatocellular carcinoma (HCC) is a major global health challenge, ranking as the sixth most frequently diagnosed cancer and the third leading cause of cancerrelated mortality worldwide^1^. The incidence and mortality rates of HCC remain alarmingly high, driven by a complex interplay of etiological factors including chronic hepatitis B and C viral infections, alcohol-associated liver disease, and increasingly, metabolic dysfunction-associated steatotic liver disease^2,3^. Despite widespread implementation of vaccination programs and direct-acting antiviral therapies that have begun to mitigate virus-related HCC, the absolute burden of the disease continues to rise globally due to aging populations and the escalating prevalence of metabolic syndrome^3^. Because HCC is frequently asymptomatic in its early stages, a substantial proportion of patients are diagnosed with advanced, unresectable disease, for which curative interventions such as surgical resection and liver transplantation are no longer viable^2^. Consequently, improving systemic therapeutic strategies and identifying robust prognostic markers are paramount priorities in contemporary hepatology and oncology.

The therapeutic landscape for advanced hepatocellular carcinoma has undergone a paradigm shift over the past decade, transitioning from a heavy reliance on single-agent tyrosine kinase inhibitors to the widespread adoption of potent combination immunotherapies^4,5^. For many years, sorafenib remained the sole approved first-line systemic therapy, offering modest survival benefits^4^. However, the advent of immune checkpoint inhibitors has profoundly altered clinical practice. The landmark success of the combination of atezolizumab, a programmed death-ligand 1 (PD-L1) inhibitor, and bevacizumab, a vascular endothelial growth factor (VEGF) inhibitor, established a new standard of care by demonstrating unprecedented improvements in overall and progression-free survival^5^. More recently, dual immune checkpoint blockade regimens, such as the combination of the cytotoxic T-lymphocyte-associated protein 4 (CTLA4) inhibitor tremelimumab and the PD-L1 inhibitor durvalumab, have further expanded the first-line armamentarium^6^. Parallel to these systemic advances, there is growing interest in integrating systemic therapies with locoregional treatments, such as transarterial chemoembolization, to maximize therapeutic efficacy and potentially downstage tumors for curative intent^7^.

As the clinical management of hepatocellular carcinoma evolves at an unprecedented pace, the volume of scientific literature dedicated to HCC treatment and prognosis is expanding exponentially. This rapid proliferation of research output presents a formidable challenge for researchers, clinicians, and policymakers striving to keep abreast of emerging trends, shifting priorities, and novel therapeutic paradigms. Traditional approaches to synthesizing this vast corpus of knowledge, such as narrative or systematic reviews, are inherently limited by human processing capacity and often struggle to capture the dynamic, real-time shifts in global research focus^8^. While conventional bibliometric analyses have been employed to map the scientific landscape, they typically rely on superficial metadata, such as predefined keywords or co-citation networks, which lack semantic granularity and may fail to accurately reflect the nuanced conceptual evolution of the field. Consequently, there is a critical need for advanced analytical methodologies capable of systematically distilling high-dimensional textual data into interpretable research themes and tracking their temporal trajectories.

Recent breakthroughs in natural language processing and the development of sophisticated large language models offer a transformative solution to the challenges of large-scale literature synthesis^8^. Unlike traditional bibliometric tools, large language models can perform deep semantic analysis of unstructured scientific texts, extracting precise, context-aware concepts that capture specific interventions, biological mechanisms, and clinical methodologies. By applying these advanced computational techniques to highly cited publications, it becomes possible to rigorously quantify thematic shifts and identify the specific research domains that are gaining or losing momentum within a given discipline. In the context of hepatocellular carcinoma, applying artificial intelligence-driven concept extraction to recent, high-impact literature could illuminate the subtle transitions in scientific focus—for example, the exact nature of the shift from broad immunotherapy concepts to specific combination regimens or the emerging emphasis on novel prognostic biomarkers. Such data-driven insights are essential for defining current frontiers, guiding future research investments, and optimizing clinical guidelines.

Therefore, the objective of this study was to characterize the evolving research emphasis within the domain of hepatocellular carcinoma treatment and prognosis by conducting a comparative bibliometric analysis of highly influential publications from two consecutive recent cohorts. By leveraging large language models for precise concept extraction and thematic normalization, we aimed to systematically quantify year-over-year frequency shifts in canonical research topics and broader parent themes among the most impactful papers. Ultimately, this approach was designed to map the shifting thematic landscape of the field and objectively delineate the most critical emerging trends shaping the contemporary management of hepatocellular carcinoma.

## Methods

### Study design and data source

This study employed a retrospective bibliometric design to characterize how research emphasis within hepatocellular carcinoma treatment and prognosis evolved between two recent publication cohorts. Bibliographic records were drawn from OpenAlex^9^ (topic T10073, Hepatocellular Carcinoma Treatment and Prognosis) and organized into the Class of 2025 (papers published in 2023) and the Class of 2026 (papers published in 2024). For each cohort, the fifty most influential on-topic articles were identified using an eighteen-month post-publication citation metric, yielding a corpus of one hundred papers for downstream concept analysis. The analytical workflow proceeded sequentially from exact citation ranking and large language model (LLM) topical screening, through automated concept extraction and cross-cohort normalization, to descriptive frequency comparison, informativeness filtering, and visualization. All primary outputs from this analysis are archived for independent reproduction (Figshare DOI: 10.6084/m9.figshare.32902769).

### Bibliographic retrieval and cohort definition

Bibliographic metadata were retrieved via the OpenAlex application programming interface^9^. Retrieval was restricted to works classified as articles within topic T10073 and to publication dates spanning 1 January 2023 through 31 December 2023 for the Class of 2025 and 1 January 2024 through 31 December 2024 for the Class of 2026. Review-type works were excluded by retaining only OpenAlex type article. At the time of analysis, the eligible article pools comprised 8,039 works for 2023 and 8,598 works for 2024. Only papers for which the full eighteen-month post-publication observation period had elapsed were considered eligible for ranking, ensuring that early citation impact could be measured on a comparable temporal footing across cohorts.

### Eighteen-month citation ranking

To assess early scientific influence, each paper was scored by the number of citing works whose publication dates fell within eighteen calendar months after the target paper’s publication date, as early citations are recognized predictors of long-term impact^10^. Because OpenAlex does not provide citation-event timestamps, this metric counts citations attributable to papers published within the window rather than citations recorded within the window. Rankings were computed with an exact threshold-expansion algorithm designed to recover the true top *N* papers by eighteen-month citation count without exhaustively scoring every work in the topic. The algorithm first retrieved an initial candidate set ranked by lifetime citations, computed exact eighteen-month counts for those candidates, and derived a provisional citation threshold from the *N* -th ranked paper in that set. All works whose lifetime citation counts met or exceeded this threshold were then scored, and the final leaderboard was sorted by exact eighteen-month counts. For each cohort, the target retention size was fifty papers; an initial candidate multiplier of 1.5 was applied when sizing internal search pools for the ranking step.

### Topical relevance screening

Because OpenAlex topic assignments can include peripherally related works, a secondary topical relevance screen was applied to the citation-ranked candidate pool. For each cohort, the one hundred highest-ranked papers by eighteen-month citations were submitted for classification (a pool size equal to twice the final target of fifty). Each paper’s title and abstract were presented to an LLM classifier (Claude Sonnet 4.6, anthropic/claude-sonnet-4.6), which returned a binary judgment of whether the study substantively addressed hepatocellular carcinoma treatment and prognosis^11,12^. Review articles were classified as on-topic when the review substantially covered the subject area. From the evaluated pool, the fifty on-topic papers with the highest eighteen-month citation counts were retained for each cohort. In each cohort, one hundred papers were evaluated; fifty-eight (Class of 2025) and sixty-five (Class of 2026) were classified as on-topic, and the final retained sets comprised the fifty highest-ranked on-topic papers per cohort.

### Concept extraction

To identify the research themes represented in the retained corpora, an LLM was used to extract concise, paper-specific concepts from each title and abstract^13^. Concept extraction was performed with Claude Sonnet 4.6 (anthropic/claudesonnet-4.6). The model was instructed to return informative concepts reflecting specific methods, technologies, named biological or clinical entities, defined sub-problems, or distinct intervention themes, while avoiding generic field-level vocabulary. A maximum of thirty concepts was requested per paper. Concepts were collected separately for each cohort before cross-cohort integration.

### Cross-cohort concept normalization

Raw extracted concepts from both cohorts were pooled and harmonized in a two-stage normalization procedure applied across all one hundred retained papers. In the first stage, 1,543 unique concept strings were grouped into 1,137 canonical concepts. Candidate synonyms were batched using embedding-based cosine similarity (Gemini Embedding 001, google/gemini-embedding-001) with a similarity threshold of 0.78, and each batch was consolidated by an LLM (DeepSeek V4 Pro, deepseek/deepseek-v4-pro) into a single canonical label^14^. In the second stage, canonical concepts were assigned to fifty-seven higher-level parent themes using the same normalization model, producing a two-tier taxonomy suitable for both fine-grained and thematic comparisons. Each paper was annotated with its normalized canonical and parent labels. When computing concept frequencies, each concept was counted at most once per paper regardless of how many times it appeared in the extraction output.

### Frequency comparison and trend metrics

Year-over-year concept prevalence was compared using cohort-normalized frequencies, defined as the number of papers in a cohort mentioning a given concept divided by the cohort size (fifty), in accordance with established bibliometric trend analysis methodologies^15^. For each concept, descriptive change metrics were computed between the Class of 2025 and the Class of 2026, including the arithmetic difference in normalized frequency, fold change, and log_2 fold change. A small constant (1 × 10 ¹²) was added to both normalized frequencies when computing fold change to avoid division by zero. These analyses were exploratory and descriptive; no formal hypothesis tests, confidence intervals, or corrections for multiple comparisons were applied. Trend figures highlighted concepts with the largest positive and negative frequency changes (ten per direction for diverging-bar and scatter plots) and the fifteen concepts with the highest Class of 2026 normalized frequency for grouped-bar displays at each taxonomy level.

### Informative concept filtering

To reduce visual and interpretive noise from generic or topic-level labels, an additional LLM review assessed whether each candidate concept conveyed information beyond what is already implied by the study topic. Classification used Claude Sonnet 5 (claude-sonnet-5). At the canonical level, the sixty concepts with the highest Class of 2026 normalized frequency were evaluated; at the parent-theme level, all fifty-seven parent themes were evaluated. Concepts classified as non-informative were excluded from trend figures and word-cloud visualizations. Informative concepts were retained for downstream display without re-estimating frequency statistics.

### Visualization

Word clouds were generated separately for each cohort using the fifty retained papers ranked by eighteen-month citations. Word size reflected the number of papers in the cohort mentioning each informative canonical concept or parent theme, with a display limit of thirty terms per cloud. Trend comparisons were rendered as diverging bar charts, scatter plots, and grouped bar charts using Matplotlib^16^. A topic-evolution summary graphic displayed the ten canonical concepts with the highest Class of 2026 normalized frequency among informative terms. Color palettes followed default Matplotlib categorical schemes.

### Software and reproducibility

All computations were performed in Python (version 3.11 or later) using pandas 3.0.3, NumPy 2.4.4^17^, Matplotlib 3.11.0^16^, WordCloud 1.9.6, and the OpenAI Python client 2.36.0 for LLM API access. Bibliographic queries were executed against the OpenAlex REST API in July 2026. LLM tasks used the models specified above: Claude Sonnet 4.6 for topical screening and concept extraction, DeepSeek V4 Pro for canonical and parent-theme normalization, Gemini Embedding 001 for similarity-based batching, and Claude Sonnet 5 for informativeness filtering. Complete ranked paper lists, extracted and normalized concepts, frequency tables, and figure-generation inputs are available in the archived dataset (DOI: 10.6084/m9.figshare.32902769).

## Results

### Composition of the influential-paper corpora

Analysis of OpenAlex topic T10073 (Hepatocellular Carcinoma Treatment and Prognosis) identified 8,039 eligible articles published in 2023 and 8,598 published in 2024. For each publication year, the one hundred highest-ranked papers by eighteen-month citation count were evaluated for topical relevance, and the fifty on-topic papers with the greatest early citation impact were retained as the Class of 2025 (2023 publications) and Class of 2026 (2024 publications) corpora. Among the evaluated pools, fifty-eight and sixty-five papers, respectively, were classified as on-topic. In the Class of 2025, eighteen-month citation counts among retained papers ranged from 504 for the top-ranked AASLD practice guidance on hepatocellular carcinoma prevention, diagnosis, and treatment to 23 for the fiftieth-ranked paper (median, 30 citations). In the Class of 2026, counts ranged from 197 for a precision-treatment review in advanced hepatocellular carcinoma to 27 for the fiftieth-ranked paper (median, 38 citations). Representative high-impact 2024 publications included four-year overall survival data from the phase III HIMALAYA study of tremelimumab plus durvalumab, an ASCO guideline update on systemic therapy for advanced hepatocellular carcinoma, and first-line results from CheckMate 9DW comparing nivolumab plus ipilimumab with lenvatinib or sorafenib. The complete ranked lists of the top 50 on-topic papers for each cohort are provided in Supplementary Tables S1^18–67^ and S2^68–117^.

### Thematic landscape of Class of 2026 publications

Word clouds generated from the fifty highest-ranked Class of 2026 papers by eighteen-month citation count characterize the dominant scientific themes of the most influential recent publications at the parent-theme level (Figure 1) and canonical-topic level (Figure 2). At the parent-theme level, the most frequently represented categories were Immunotherapy and immune biology (26 paper mentions), HCC biology and tumor microenvironment (13 mentions), and Cancer Immunotherapy (13 mentions). At the canonical-topic level, immune checkpoint inhibitor therapy was the single most frequent concept (24 mentions), followed by PD-1/PD-L1 inhibitor (20 mentions), sorafenib (19 mentions), and unresectable hepatocellular carcinoma, combination immunotherapy, and hepatectomy (10 mentions each).

**Figure 1.**
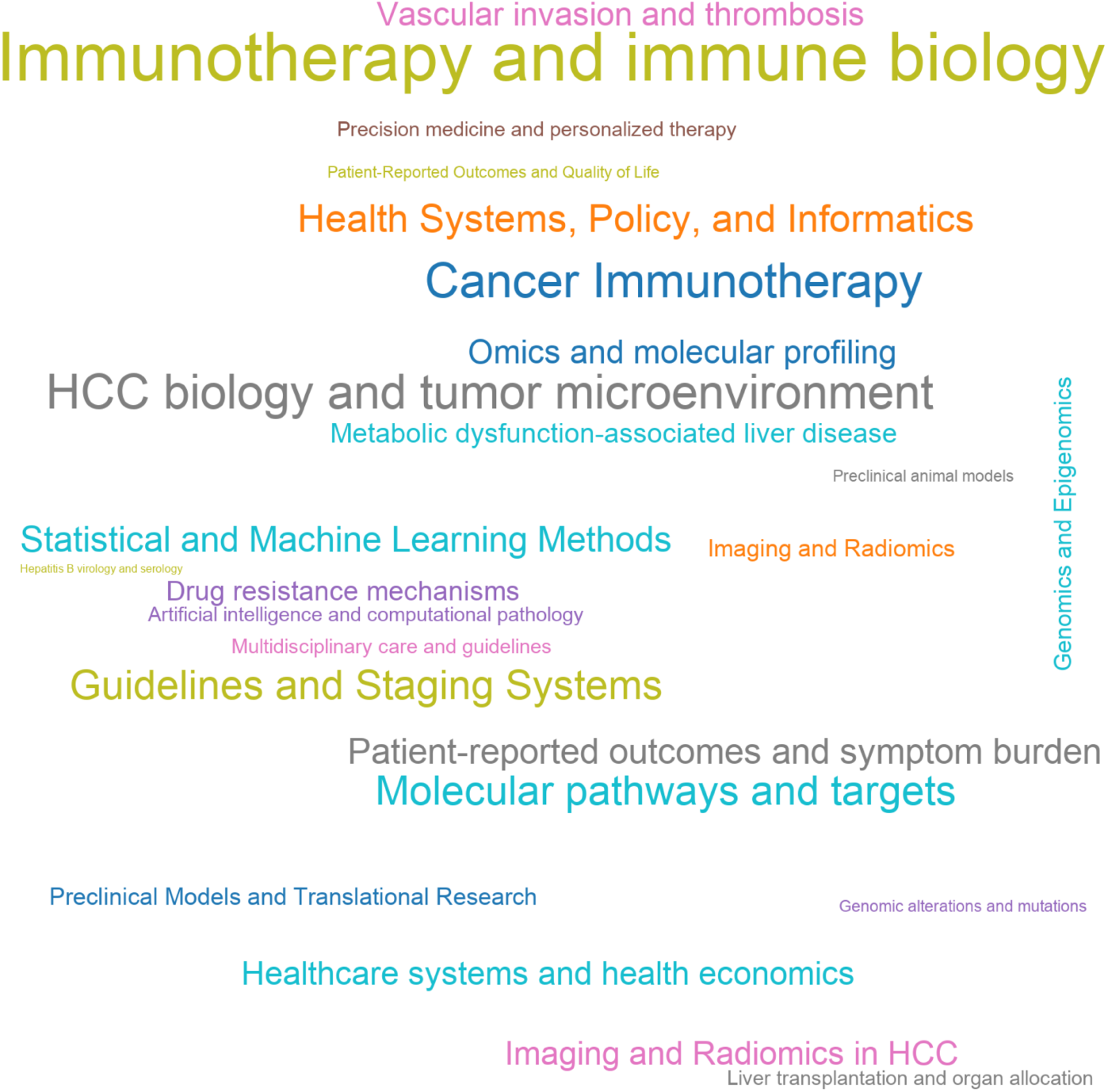
Word cloud of parent themes across the 50 most-cited Class of 2026 papers. Word size reflects the number of Class of 2026 papers mentioning each informative parent theme (maximum thirty terms displayed). Abbreviations: HCC, hepatocellular carcinoma.

**Figure 2.**
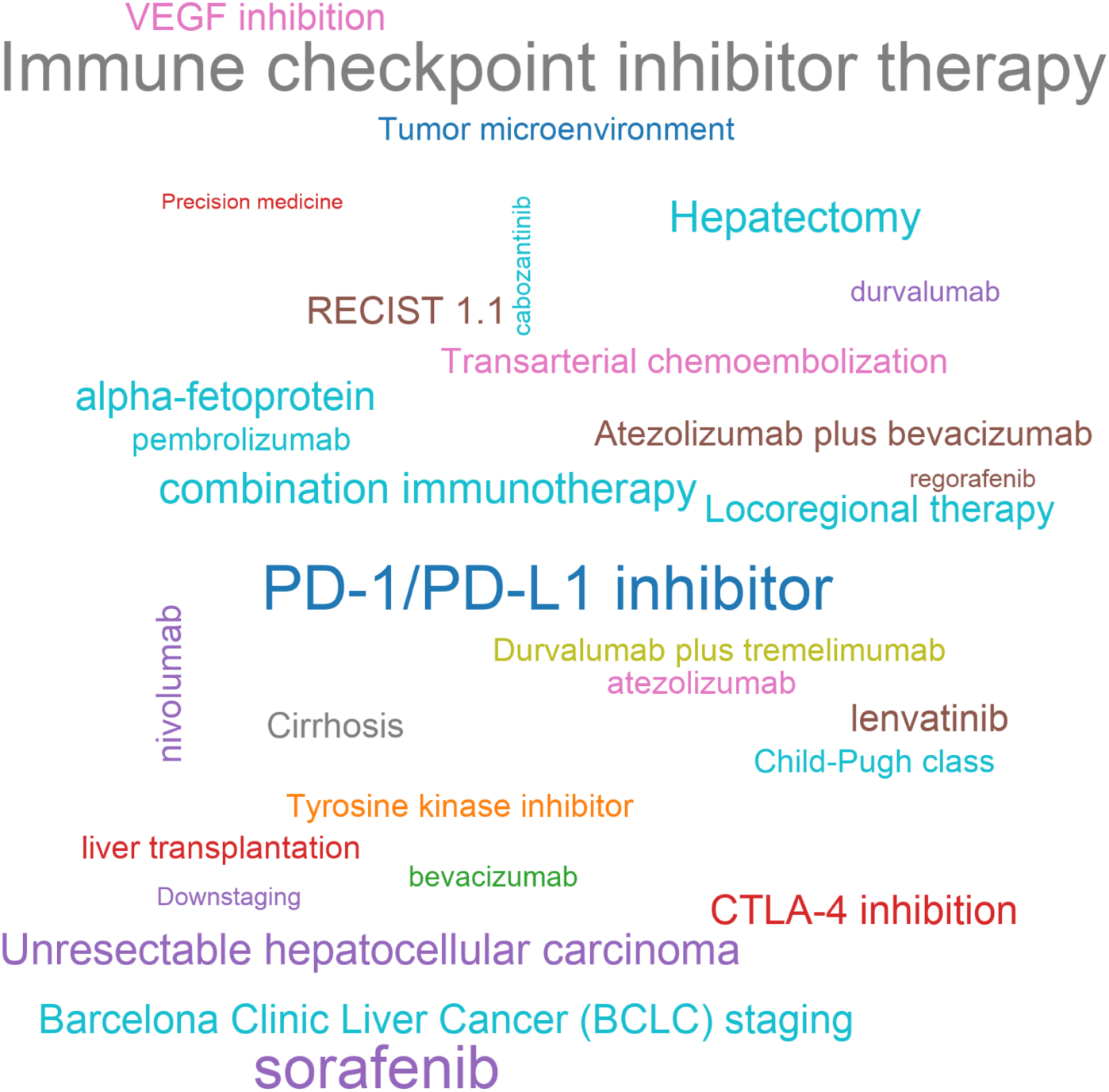
Word cloud of canonical topics across the 50 most-cited Class of 2026 papers. Word size reflects the number of Class of 2026 papers mentioning each informative canonical topic (maximum thirty terms displayed). Abbreviations: HCC, hepatocellular carcinoma; PD-1, programmed cell death protein 1; PDL1, programmed death-ligand 1; CTLA-4, cytotoxic T-lymphocyte-associated protein 4; BCLC, Barcelona Clinic Liver Cancer; TACE, transarterial chemoembolization; TKI, tyrosine kinase inhibitor; VEGF, vascular endothelial growth factor; RECIST, Response Evaluation Criteria in Solid Tumors.

### Parent-theme frequency shifts between cohorts

Comparison of normalized concept frequencies between the Class of 2025 and Class of 2026 cohorts revealed systematic shifts in the thematic composition of highly cited hepatocellular carcinoma publications (Figure 3). The diverging bar chart (Figure 3a) highlighted parent themes with the largest cohort differences. Among themes with the largest increases in normalized frequency in the Class of 2026 cohort, Imaging and Radiomics in HCC showed the greatest gain (frequency difference, +0.12; papers containing the theme, 1 in Class of 2025 versus 7 in Class of 2026), followed by Health Systems, Policy, and Informatics (+0.10; 4 versus 9), Molecular pathways and targets (+0.08; 6 versus 10), Healthcare systems and health economics (+0.08; 3 versus 7), Drug resistance mechanisms (+0.08; 1 versus 5), and Cancer Immunotherapy (+0.06; 10 versus 13). Additional themes with positive shifts included HCC biology and tumor microenvironment (+0.04; 11 versus 13), Imaging and Radiomics (+0.04; 2 versus 4), and Preclinical Models and Translational Research (+0.04; 2 versus 4).

**Figure 3.**
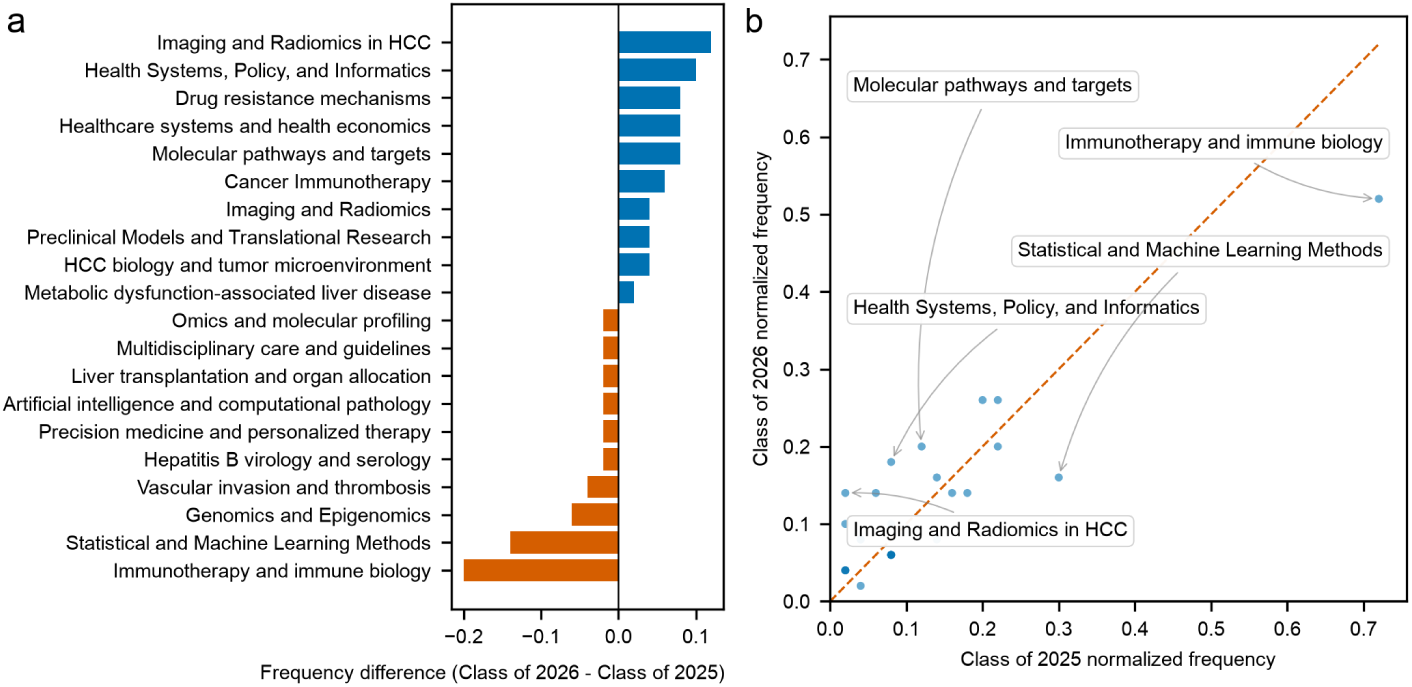
Cohort comparison of concept frequencies at the parent-theme level. Panel a, horizontal diverging bar chart of normalized frequency difference (Class of 2026 minus Class of 2025) for parent themes with the largest absolute shifts; blue bars indicate positive differences and orange bars indicate negative differences. Panel b, scatter plot with Class of 2025 normalized frequency on the x-axis and Class of 2026 normalized frequency on the y-axis; the dashed diagonal line denotes equal frequency between cohorts; selected outlier themes are annotated. Abbreviations: HCC, hepatocellular carcinoma.

Among parent themes with the largest decreases (Figure 3a), Immunotherapy and immune biology declined most markedly (frequency difference, -0.20; 36 versus 26), followed by Statistical and Machine Learning Methods (-0.14; 15 versus 8), Genomics and Epigenomics (-0.06; 7 versus 4), and Vascular invasion and thrombosis (-0.04; 9 versus 7). Smaller negative shifts were observed for Precision medicine and personalized therapy (-0.02; 4 versus 3), Liver transplantation and organ allocation (-0.02; 4 versus 3), Artificial intelligence and computational pathology (-0.02; 4 versus 3), and Hepatitis B virology and serology (-0.02; 2 versus 1). The scatter plot (Figure 3b) showed that most parent themes clustered near the diagonal of equal normalized frequency, with annotated outliers corresponding to the themes with the largest absolute frequency differences described above. Complete parent-theme counts and change metrics for all fifty-seven themes are reported in Supplementary Table S3 (Supplementary Table S3).

### Prevalence of leading parent themes in the Class of 2026 cohort

Among the fifteen informative parent themes with the highest normalized frequency in the Class of 2026 cohort (Figure 4), Immunotherapy and immune biology ranked highest (normalized frequency, 0.52), although this theme was more prevalent in the Class of 2025 cohort (0.72). HCC biology and tumor microenvironment and Cancer Immunotherapy co-ranked next (0.26 each), with both showing higher representation in the Class of 2026 than in the Class of 2025 (0.22 and 0.20, respectively). Guidelines and Staging Systems (0.20 versus 0.22) and Molecular pathways and targets (0.20 versus 0.12) followed. Health Systems, Policy, and Informatics (0.18 versus 0.08), Healthcare systems and health economics (0.14 versus 0.06), Imaging and Radiomics in HCC (0.14 versus 0.02), and Drug resistance mechanisms (0.10 versus 0.02) showed markedly higher normalized frequency in the Class of 2026 cohort. By contrast, Statistical and Machine Learning Methods (0.16 versus 0.30), Vascular invasion and thrombosis (0.14 versus 0.18), and Omics and molecular profiling (0.14 versus 0.16) were more represented in the Class of 2025 cohort.

**Figure 4.**
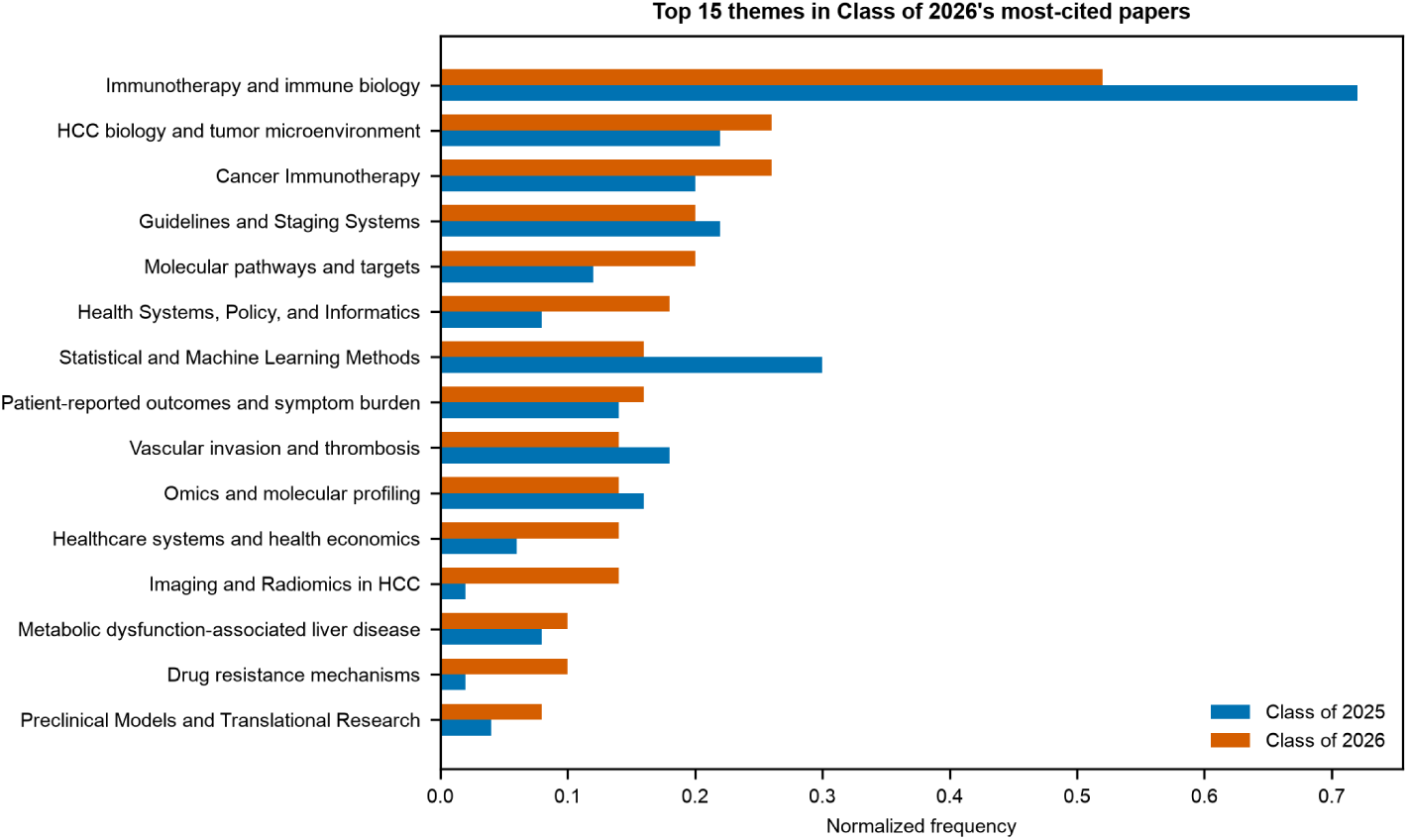
Normalized frequencies of the 15 most prevalent parent themes in the Class of 2026 cohort. Horizontal grouped bars show normalized frequency for the Class of 2025 (blue) and Class of 2026 (orange) for each listed parent theme; themes are ordered by Class of 2026 prevalence. Abbreviations: HCC, hepatocellular carcinoma.

### Canonical-topic frequency shifts between cohorts

At the canonical-topic level, comparison of normalized concept frequencies between cohorts revealed a shift from broad therapeutic labels toward specific immunotherapy agents and regimens (Figure 5). The diverging bar chart (Figure 5a) showed that CTLA-4 inhibition exhibited the largest increase in normalized frequency (frequency difference, +0.14; papers containing the topic, 2 in Class of 2025 versus 9 in Class of 2026). Durvalumab plus tremelimumab showed the next-largest gain (+0.12; 0 versus 6), followed by pembrolizumab (+0.10; 1 versus 6), nivolumab (+0.08; 3 versus 7), durvalumab (+0.08; 1 versus 5), and sorafenib (+0.06; 16 versus 19). Other canonical topics with notable increases included bridging therapy (+0.06; 0 versus 3), VEGF signaling pathway (+0.06; 0 versus 3), HCC management (+0.06; 0 versus 3), and Competing risks analysis (+0.06; 0 versus 3).

**Figure 5.**
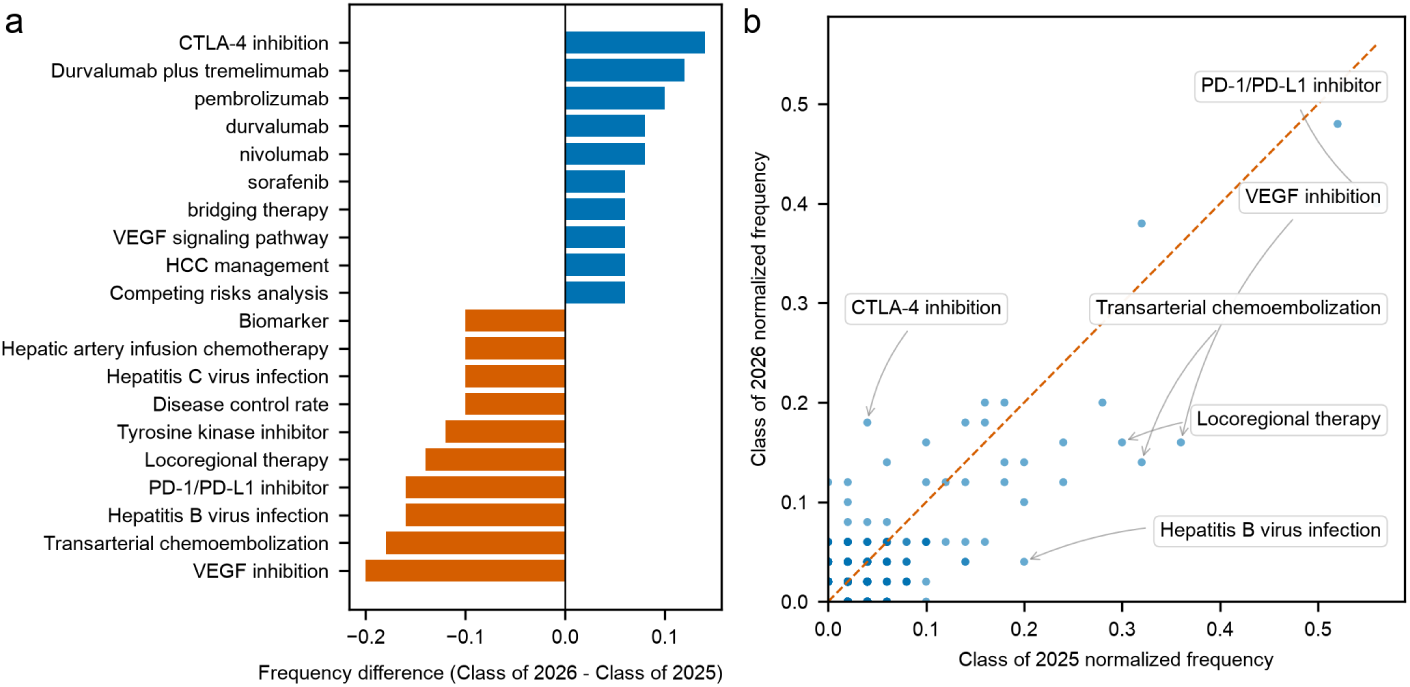
Cohort comparison of concept frequencies at the canonical-topic level. Panel a, horizontal diverging bar chart of normalized frequency difference (Class of 2026 minus Class of 2025) for canonical topics with the largest absolute shifts; blue bars indicate positive differences and orange bars indicate negative differences. Panel b, scatter plot with Class of 2025 normalized frequency on the x-axis and Class of 2026 normalized frequency on the y-axis; the dashed diagonal line denotes equal frequency between cohorts; selected outlier topics are annotated. Abbreviations: CTLA-4, cytotoxic T-lymphocyte-associated protein 4; PD-1, programmed cell death protein 1; PD-L1, programmed death-ligand 1; VEGF, vascular endothelial growth factor; TACE, transarterial chemoembolization.

Canonical topics with the largest decreases (Figure 5a) included VEGF inhibition (frequency difference, -0.20; 18 versus 8), transarterial chemoembolization (-0.18; 16 versus 7), PD-1/PD-L1 inhibitor (-0.16; 28 versus 20), Hepatitis B virus infection (-0.16; 10 versus 2), locoregional therapy (-0.14; 15 versus 8), and tyrosine kinase inhibitor (-0.12; 12 versus 6). Hepatitis C virus infection (-0.10; 8 versus 3) and biomarker (-0.10; 7 versus 2) also appeared among declining terms. As in the parent-theme analysis, the scatter plot (Figure 5b) placed most canonical topics near the diagonal, with annotated outliers marking the topics with the largest absolute frequency differences. Full canonical-topic metrics for all 1,137 harmonized concepts are listed in Supplementary Table S4 (Supplementary Table S4).

### Prevalence of leading canonical topics in the Class of 2026 cohort

Among the fifteen canonical topics with the highest normalized frequency in the Class of 2026 cohort (Figure 6), immune checkpoint inhibitor therapy ranked highest (0.48), followed by PD-1/PD-L1 inhibitor (0.40) and sorafenib (0.38). Hepatectomy, unresectable hepatocellular carcinoma, and combination immunotherapy each showed normalized frequency of 0.20. Barcelona Clinic Liver Cancer (BCLC) staging, CTLA-4 inhibition, and alpha-fetoprotein each reached 0.18. Locoregional therapy, RECIST version 1.1, VEGF inhibition, and lenvatinib were represented at 0.16. Cirrhosis and nivolumab each showed normalized frequency of 0.14 (Class of 2025, 0.18 and 0.06, respectively). Compared with the Class of 2025 cohort, sorafenib, CTLA-4 inhibition, hepatectomy, RECIST version 1.1, and nivolumab showed higher normalized frequency in the Class of 2026 cohort, whereas PD-1/PD-L1 inhibitor, VEGF inhibition, locoregional therapy, lenvatinib, cirrhosis, and unresectable hepatocellular carcinoma were more prevalent in the Class of 2025 cohort.

**Figure 6.**
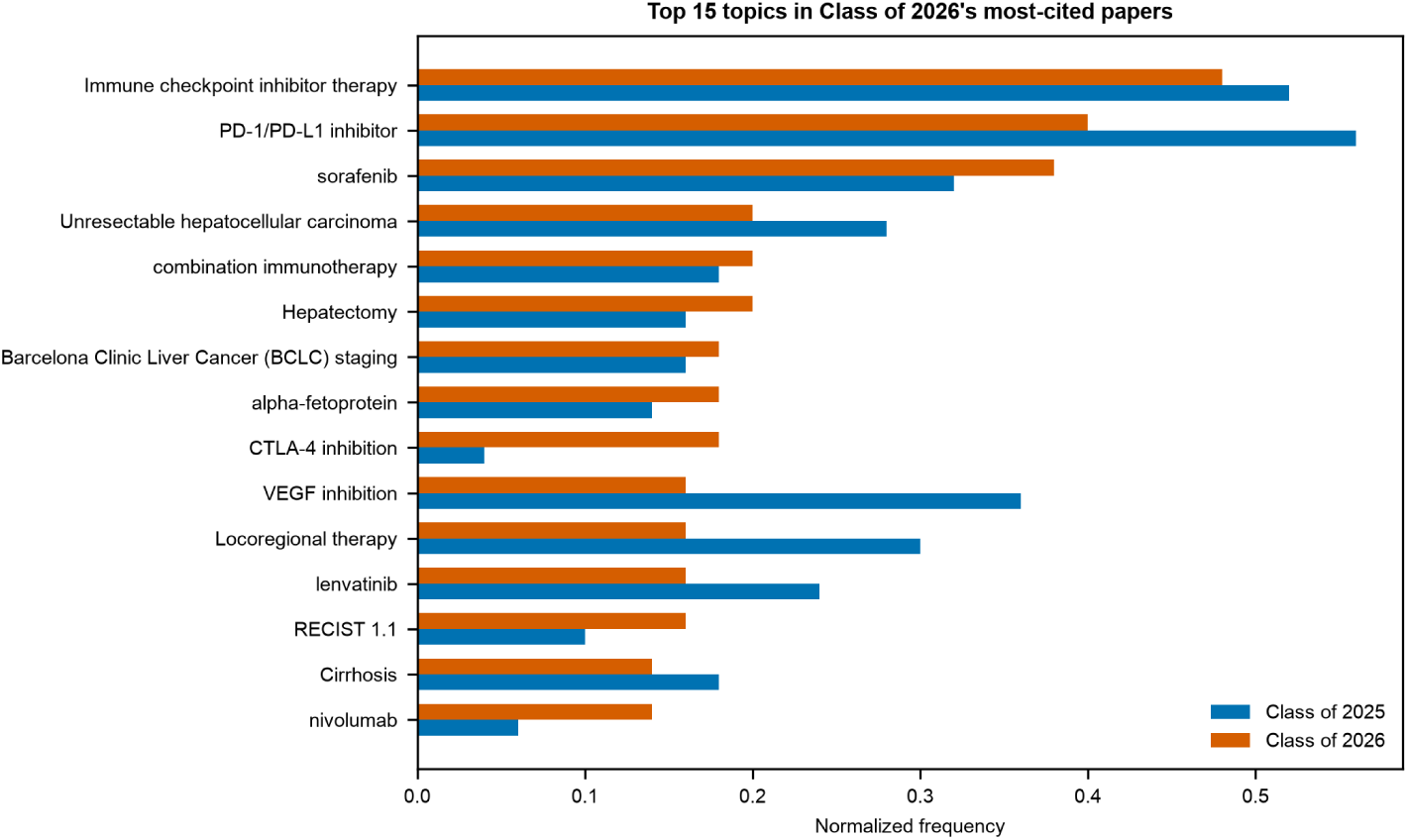
Normalized frequencies of the 15 most prevalent canonical topics in the Class of 2026 cohort. Horizontal grouped bars show normalized frequency for the Class of 2025 (blue) and Class of 2026 (orange) for each listed canonical topic; topics are ordered by Class of 2026 prevalence. Abbreviations: PD-1, programmed cell death protein 1; PD-L1, programmed death-ligand 1; CTLA4, cytotoxic T-lymphocyte-associated protein 4; BCLC, Barcelona Clinic Liver Cancer; VEGF, vascular endothelial growth factor; RECIST, Response Evaluation Criteria in Solid Tumors.

## Discussion

This study deployed a novel large language model-driven bibliometric approach to rapidly characterize the evolving landscape of hepatocellular carcinoma treatment and prognosis by analyzing the most highly cited literature published in 2023 and 2024. Our findings reveal a decisive maturation in the field of systemic therapy, characterized by a transition from broad investigations of single-agent immune checkpoint inhibitors and tyrosine kinase inhibitors toward specific, potent combination immunotherapies, particularly those incorporating dual checkpoint blockade. Concurrently, we identified a marked ascendance in research emphasis on imaging, radiomics, and health systems informatics, alongside a relative contraction in the prominence of traditional locoregional interventions such as transarterial chemoembolization^118,119^.

The thematic pivot toward specific immunotherapeutic regimens, notably CTLA-4 inhibition in combination with PD-1 or PD-L1 blockade, directly reflects recent landmark successes in late-stage clinical trials that have reshaped first-line treatment algorithms for unresectable hepatocellular carcinoma. The profound increases in the normalized frequencies of terms such as “CTLA-4 inhibition,” “durvalumab plus tremelimumab,” and “nivolumab” correspond to the maturation of survival data from pivotal studies. For example, the phase III HIMALAYA trial established the long-term overall survival benefit of the single tremelimumab regular interval durvalumab (STRIDE) regimen over traditional tyrosine kinase inhibitors, fundamentally altering the therapeutic landscape^69^. Similarly, the robust citation impact surrounding the CheckMate 9DW trial^120^ underscores the clinical community’s intense focus on nivolumab plus ipilimumab as a formidable therapeutic option capable of eliciting deep and durable responses. The transition away from generic terms like “PD-1/PD-L1 inhibitor,” “tyrosine kinase inhibitor,” and “VEGF inhibition” in the 2024 cohort does not indicate a waning interest in these biological pathways. Rather, it signifies a field moving beyond initial proof-of-concept monotherapies toward the rigorous refinement of specific, synergistic combinations designed to overcome both primary and acquired resistance in the tumor microenvironment^121^.

These findings contextualize and substantially extend upon prior bibliometric analyses of hepatocellular carcinoma. Earlier retrospective mappings of the biomedical literature documented the initial surge of interest surrounding the introduction of sorafenib as the first effective systemic therapy, followed by the subsequent wave of enthusiasm for early single-agent PD-1 inhibitors^122^. Our analysis captures the leading edge of a subsequent evolutionary wave, defining a new era dominated by dual combination immunotherapy and precision patient selection. Furthermore, the rising prominence of “Imaging and Radiomics in HCC” in our dataset aligns with parallel trends observed across global oncology. Recent comprehensive reviews emphasize that artificial intelligence and radiomic features extracted from routine cross-sectional imaging—such as computed tomography and magnetic resonance imaging—are increasingly being investigated as non-invasive biomarkers for predicting microvascular invasion, molecular subclass, and response to immunotherapy^123^. Our data quantitatively confirm that this previously niche domain is now generating some of the most influential, highly cited, and rapidly disseminating publications in the field of hepatology.

A key methodological innovation of this study is the deployment of large language models for concept extraction and harmonization across publication cohorts. Traditional bibliometric techniques often rely on author-provided keywords, Medical Subject Headings, or rigid natural language processing pipelines, which can lag behind emerging terminology or struggle to distinguish nuanced clinical concepts. By leveraging advanced language models, we were able to extract highly granular, context-aware concepts from unstructured abstracts and seamlessly map them to canonical topics and parent themes. This approach facilitated a highly responsive, near real-time tracking of research dynamics, effectively capturing the immediate impact of breaking clinical trial data within an eighteen-month post-publication window. The ability to filter concepts for specific informational value further enhanced the signal-to-noise ratio, revealing the exact terminology driving current scientific discourse^124^.

The shifting thematic landscape identified here carries substantial scientific and translational implications. As the therapeutic armamentarium for unresectable hepatocellular carcinoma rapidly expands with complex, high-cost combination immunotherapies, the clinical challenge fundamentally shifts from de novo drug discovery to optimal treatment sequencing, patient stratification, and the proactive mitigation of immune-mediated adverse events. This clinical reality is directly mirrored in our data by the concurrent rise in publications focused on health systems, health economics, competing risks analysis, and informatics. The growing intersection of advanced radiomics, artificial intelligence, and health policy suggests that the next major breakthroughs in hepatocellular carcinoma may not solely involve novel pharmacological agents. Instead, they will likely emerge from the development of integrated, multidisciplinary predictive models that optimize the delivery of existing advanced therapies. By identifying which patients are most likely to benefit from dual checkpoint blockade versus combined immune and anti-angiogenic therapy, these models have the potential to maximize survival, minimize toxicity, and deliver superior value within real-world, diverse patient populations^123,125^.

The strengths of this study lie in its objective, algorithmic identification of the most impactful literature and its use of an eighteen-month citation window to capture immediate scientific influence, circumventing the biases inherent in lifetime citation counts which heavily favor older publications. However, several limitations must be acknowledged. The reliance on an eighteen-month citation metric inherently favors large, phase III clinical trials and comprehensive clinical guidelines, which rapidly accumulate citations, potentially underrepresenting fundamental basic science or preclinical research that requires a longer gestation period to be widely recognized. Additionally, our dataset was restricted to articles indexed in OpenAlex within a specific timeframe; thus, highly influential papers published outside this window or categorized under different topic clusters may have been omitted. Finally, while large language models offer unprecedented capabilities in text analysis, their extraction and normalization outputs remain susceptible to subtle biases or hallucinated alignments, although our rigorous similarity-based batching and multi-tier taxonomy were explicitly designed to mitigate these risks^126^.

Future research directions should build upon these bibliometric insights by prospectively integrating real-time data from clinical trial registries and preprint servers to forecast emerging therapeutic trends before they manifest in the peerreviewed literature. Additionally, subsequent analyses should investigate the geographic and institutional networks driving these thematic shifts to identify global disparities in research focus and clinical trial access. In the clinical domain, the strong signal favoring radiomics suggests an urgent need for largescale, prospective validation of artificial intelligence-driven imaging biomarkers across multi-center cohorts to effectively guide the selection of dual checkpoint blockade versus combination immunotherapy and anti-angiogenic regimens. Furthermore, qualitative studies exploring how clinicians incorporate these rapidly evolving treatment paradigms into real-world practice will be essential for identifying and overcoming barriers to implementation.

In conclusion, this bibliometric analysis demonstrates that the most influential research in hepatocellular carcinoma is currently defined by the optimization of complex systemic immunotherapies and the parallel pursuit of non-invasive predictive technologies. The rapid displacement of broad therapeutic concepts by specific, potent combination regimens such as durvalumab plus tremelimumab and nivolumab plus ipilimumab highlights a field experiencing profound clinical acceleration and transformation. Moving forward, the strategic integration of computational pathology, radiomics, and health economics will be absolutely essential to translate these ongoing therapeutic advances into durable, equitable, and widespread improvements in patient outcomes globally.

## Acknowledgments

We extend our sincere gratitude to the OpenAlex initiative for supplying the foundational bibliometric dataset that made this analysis possible. We also thank the developers and communities behind the open-source software tools and models employed during data processing and evaluation, whose efforts were indispensable to our study.

## Data availability

The data generated and analyzed in this study are publicly accessible on Figshare at https://doi.org/10.6084/m9.figshare.32902769. The complete source code and analytical pipelines are maintained on GitHub at https://github.com/pepkio/pri-top-papers. Furthermore, an interactive web interface for exploring our findings is provided at https://pri.pepkio.com.

## Supplementary Tables

**Supplementary Table S1.** Ranked list of the fifty Class of 2025 (2023) papers retained after eighteen-month citation ranking and topical screening, including rank, eighteen-month citation count, topical relevance classification, article title, journal, publication date, and DOI.

**Supplementary Table S2.** Ranked list of the fifty Class of 2026 (2024) papers retained after eighteen-month citation ranking and topical screening, including rank, eighteen-month citation count, topical relevance classification, article title, journal, publication date, and DOI.

**Supplementary Table S3.** Parent-theme frequency comparison across cohorts, including parent-theme name, paper counts in 2023 and 2024, normalized frequencies, frequency difference, fold change, and log_2 fold change.

**Supplementary Table S4.** Canonical-topic frequency comparison across cohorts, including canonical topic name, paper counts in 2023 and 2024, normalized frequencies, frequency difference, fold change, and log_2 fold change.

